# Impact of genetic counselling strategy on diagnostic yield and workload for genome sequencing-based tumour diagnostics

**DOI:** 10.1101/2023.07.11.23291187

**Authors:** Roelof Koster, Luuk J. Schipper, Noor A. A. Giesbertz, Daphne van Beek, Matías Mendeville, Kris G. Samsom, Efraim H. Rosenberg, Frans B.L. Hogervorst, Paul Roepman, Mirjam C. Boelens, Linda J.W. Bosch, Jose G. van den Berg, Gerrit A. Meijer, Emile E. Voest, Edwin Cuppen, Marielle W.G. Ruijs, Tom van Wezel, Lizet van der Kolk, Kim Monkhorst

**Affiliations:** The Netherlands Cancer Institute, Amsterdam, The Netherlands; Hartwig Medical Foundation, Amsterdam, The Netherlands

**Keywords:** Genome Sequencing, validation, cancer genetics, molecular diagnostics, Germline

## Abstract

**Purpose:** Genome sequencing (GS) enables comprehensive molecular analysis of tumours and identification of hereditary cancer predisposition. According to guidelines, directly determining pathogenic germline variants (PGVs) requires pre-test genetic counselling, which is cost-ineffective. Referral for genetic counselling based on tumour variants alone could miss relevant PGVs and/or result in unnecessary referrals.

**Methods:** We validated GS for detection of germline variants and simulated three strategies using paired tumour-normal genome sequencing data of 937 metastatic patients. In strategy-1 genetic counselling prior to tumour testing allowed direct PGV analysis. In strategy-2 and -3, germline testing and referral for post-test genetic counselling is based on tumour variants using Dutch (strategy-2) or ESMO-PMWG (strategy-3) guidelines.

**Results:** In strategy-1, PGVs would be detected in 50 patients (number-needed-to counsel; NTC=18.7). In strategy-2, 86 patients would have been referred for genetic counselling and 43 would have PGVs (NTC=2). In strategy-3, 94 patients would have been referred for genetic counselling and 32 would have PGVs (NTC=2.9). Hence, 43 and 62 patients, respectively, were unnecessarily referred based on a somatic variant.

**Conclusion:** Both post-tumour test counselling strategies (2 and 3) had significantly lower NTC, and strategy-2 had the highest PGV yield. Combining pre-tumour test mainstreaming and post-tumour test counselling may maximize the clinically relevant PGV yield and minimize unnecessary referrals.

## Introduction

The druggable genome is rapidly expanding and requires continuously adaptation of molecular diagnostic panels for therapy selection based on the latest actionable genomic alterations. Therefore, broad panels like TSO500, PGDx elio, MSK-IMPACT and genome sequencing are essential developments to analyse the tumours of (metastatic) cancer patients^1–5^. Through the increasing use of these broad molecular diagnostic panels, pathogenic germline variants (PGVs) are now more frequently identified following tumour-based DNA sequencing. Importantly, this includes patients that do not meet current genetic testing criteria^6–18^. Those criteria including age of onset, personal and/or family history and tumour characteristics^19–22^. For better analysis of genomic tumour data these broad tests can use ‘normal’ DNA (i.e. germline DNA; usually from blood) as a reference. With this comes the possibility of simultaneous detection of pathogenic germline variants (PGVs).

Detection of PGVs becomes increasingly important in targeted therapies. The germline status can be pivotal for therapy selection, such as for *BRCA1/2* mutated ovarian, breast, pancreas and prostate cancer patients^23–26^. Hence, the 2020 ESMO Clinical Practice Guidelines for prostate cancer recommended the consideration of *BRCA1/2* germline sequencing in all patients with metastatic prostate cancer^27^. Furthermore, PGVs signify a hereditary cancer predisposition providing valuable information for timely surveillance and preventative interventions for both the patient and at-risk family members^28–31^. Interestingly, 11% of patients with a positive finding from tumour sequencing had their PGVs detected only after presenting with a second primary cancer. Many of which would be preventable or detected earlier if the PGV had been detected with the first primary^32^, further underscoring the importance of PGV detection and secondary prevention in cancer patients. Although the importance of detecting PGVs is indisputable, at the same time, it is important to incorporate germline analysis in such a way that patients are adequately informed about possible germline findings and provide consent, before the germline is analysed. Furthermore, it’s important to note that some PGVs may not result in clinical implications for patients or their family members, because of too limited knowledge about associated risks and/or the absence of surveillance and prevention recommendations. In Europe, not all of these PGV secondary findings need to be actively analyzed and reported, as stated by the European Society of Human Genetics (ESHG)^33^. In the United States however these findings should be reported especially concerning cancer risk genes listed in the ACMG SF v3.1^34^. If a patient has chosen not to receive secondary findings from genomic sequencing, there may be no requirement to analyze or report these findings, in accordance with the American College of Medical Genetics and Genomics (ACMG) policy^35^.

Genomic tumour profiling of (metastatic) cancer patients therefore requires an integrated strategy to refer patients effectively to a clinical geneticist. Unnecessary referrals should be prevented because of possible distress for patients and the associated workload for treating physicians and/or clinical geneticists. Strategies could be based on current guidelines for putative germline variants detected with tumour DNA sequencing, such as developed by the ESMO Precision Medicine Working Group (ESMO-PMWG) and the Dutch Working Group tumour- and germline-diagnostics^3, 4, 36^. These prioritize which tumour-detected putative germline variants to follow up on. Because of the tumour-based approach relevant PGVs may be missed while somatic tumour variants in cancer predisposition genes may lead to unnecessary referrals. Another strategy is to first refer patients to genetic counsellors which then allows direct reporting of all relevant PGVs upon patients consent parallel with the tumour analysis report. This approach is cost-ineffective and poses a burden on patients. Hence, both strategies, i.e. direct germline analysis and germline analysis based on tumour variants, have their advantages and disadvantages.

In the <u>W</u>GS Implementation in standard <u>D</u>iagnostics for <u>E</u>ach cancer patient (WIDE) study^37, 38^ we performed paired tumour-normal genome sequencing on 937 metastatic cancer patients. To investigate the optimal strategy to detect hereditary predisposition we compared the PGV yield and the number-needed-to-counsel (NTC) of a simulated pre-tumour test counselling strategy with two simulated post-tumour test counselling strategies using the WIDE data.

## Methods

### Study population

Patients were included as part of the <u>W</u>GS Implementation in standard <u>D</u>iagnostics for <u>E</u>ach cancer patient (WIDE) study^37, 38^. A description of the study protocol and technical details of the genome sequencing analysis can be found elsewhere^37, 38^. In short, metastatic solid cancer patients, unselected for tumour type, age of onset and/or family history, received genome sequencing analysis in parallel with standard-of-care diagnostics to study the feasibility, validity, and clinical value of genome sequencing based tumour diagnostics in routine pathology practice.

### Genome sequencing, bioinformatics and reporting

Genome sequencing was performed at the Hartwig Medical Foundation (Amsterdam, The Netherlands). Tumour DNA was analysed with a sequencing depth of >90-110x on Illumina NovaSeq6000 platforms (Illumina, San Diego, CA, USA) according to previously described procedures ^37–39^. DNA isolated from blood was sequenced at a minimal depth of >30x and was used as a reference, allowing to discriminate somatic variants from germline DNA background variants in bioinformatic analyses. A standardized, fully open source bioinformatics pipeline that is continuously evaluated and improved when necessary^40^, was used to analyse sequencing data based on reference genome version GRCh37/hg19 (code available through **<u>github.com/hartwigmedical)</u>**. The SAGE caller (https://github.com/hartwigmedical/hmftools/blob/master/sage/), for single nucleotide variants, multi-nucleotide variants and small insertions/deletions and the GRIPPS caller (https://github.com/hartwigmedical/hmftools/tree/master/gripss) for structural variants and genomic breakpoint junctions were adjusted for improved germline variant detection. The genome sequencing report (Hartwig Medical OncoAct, Amsterdam, The Netherlands) reported all somatic variants with a (high) driver likelihood ^39^, including variants, copy number alterations, fusions in 460 genes and mutational signatures (such as tumour mutational load, homologues recombination deficiency (HRD) and microsatellite instability (MSI)) for each patient. In general variants that are present in the germline and tumour are excluded from the OncoAct genome sequencing report, except for variants in 57 genes that were solely selected because of diagnostic and/or therapeutic relevance, not necessarily for detecting predisposition to cancer (for instance: *CHEK1* for experimental PARPi treatment and *TP53* for determining clonality and for prognosis; see Supplemental Table 1, Supplemental Figure 1).

Notably, only variants in these genes that are present in both germline and tumour are included on the OncoAct genome sequencing report as variants present in the tumour, although their germline status is not explicated annotated, while variants that are present solely in the germline are excluded from the OncoAct genome sequencing report because the genome sequencing diagnostic test is focused on clinical actionability and not intended to be a germline diagnostics tool.

### Germline – panel of 49 genes & retrospective analysis

Based on available literature, recommendations and guidelines, 49 of the 57 genes (see Supplemental Table 1, Supplemental Figure 1) are established cancer predisposition genes (CPGs) ^3, 4, 19–22, 34, 36, 41–45^. CPGs are genes that can harbour germline variants that confer high or moderate risk to certain cancer types. Based on an unbiased, pseudonymized germline screening of these 49 cancer-associated genes, in paired tumour-normal genome sequencing data of 937 patients, all germline variants reported by the bioinformatical pipeline were manually reviewed by at least one Clinical Laboratory Geneticist to determine pathogenicity of reported variants (for gene reference transcripts used see Supplemental Table 1). All (likely) pathogenic variants were considered as baseline and used to determine the prevalence of PGVs in the overall cohort. For patients with a known pathogenic germline variant prior to genome sequencing analysis, clinical data were available.

### Validation of germline analysis from genome sequencing

138 patients within the WIDE study had already received genetic (germline) testing prior to genome sequencing analysis with concordant results. 28 patients had a PGV reported with both genome sequencing and genetic testing and 2 patients had a PGV reported with genome sequencing that was not covered in the gene panel at the time. One patient had a variant reported with previous genetic testing that is currently considered benign. 108 patients (including the one with the benign finding) had no relevant finding using genome sequencing. PGVs reported included single nucleotide variants, multi-nucleotide variants, indels and structural variants of which two were in exon 14 of *PMS2* (difficult to detect because of the pseudogene region).

In 2021 we implemented tumour genome sequencing in routine care and (with consent of the patients) germline variants were annotated as such on the report. We reported for 37 patients relevant PGVs that were either know prior to genome sequencing or confirmed. 29 patients had no relevant PGVs reportable with genome sequencing which was concordant with genetic panel testing performed prior. In addition, 20 DNAs from blood with a complex and/or difficult to detect germline finding from regular diagnostics (SNVs, indels and CNVs some of which are located in pseudogene region or stretch) were selected from our archive and tested using 30x genome sequencing. In these 20 samples >99,5% (median and average) of bases within the gene panel is covered with > 20 reads. All complex and/or difficult to detect variants were detected. Although with low quality scores we also detected 2 deletions in *PMS2*. Taken together, detection of pathogenic germline variants and structural variants has been validated on a total set of 224 patients with a sensitivity (re-call) of 100% (95%CI 94.7 – 100%) and a specificity of 100% (95%CI 96.6 – 100%; see Supplemental Figure 2).

### Germline analysis using three different simulated strategies

In strategy-1, germline DNA is simultaneously analysed with the genome sequencing tumour analysis and clinically relevant PGVs are reported based on a tumour type-specific gene panel (gene information and reference transcripts used see Supplemental Table 1). This tumour type-specific gene panel is based on the standard of care genetic (germline) testing gene panels used for the specific tumour type(s) as indicated^19–22^. This strategy requires that all 937 patients are counselled about the possibility of germline findings as a part of the genome sequencing test results.

In strategy-2 and strategy-3, all genome sequencing detected tumour variants in the selected 49 genes were reviewed by at least one Clinical Laboratory Geneticist to determine pathogenicity (for gene reference transcripts used see Supplemental Table 1), after they matched the adjusted Dutch guidelines^36^ and/or the recommendations of the ESMO Precision Medicine Working Group (ESMO PMWG)^3, 4^ for tumour sequencing (Supplemental Table 1). In our hospital we also wanted to detect patients heterozygous for a pathogenic variant in the MMR/Lynch genes, therefore we adjusted the Dutch guidelines for the MMR/Lynch genes and reported tumour variants in these genes for any tumour type (in the original Dutch guidelines patients are referred when a variant in MMR/Lynch genes is detected for any tumour type and MSI/dMMR is present in patients <70 years).

Next, according to the recommendations outlined in the related guideline, it is essential to refer patients for genetic counselling regarding germline analysis. After consent of the patient, germline testing can be performed to analyse if the reported pathogenic tumour variant (PTV) in the genome sequencing report is present in the germline or not. Here, we report how many of these patients had a (likely) PGV or a somatic variant as the basis for their referral for genetic counselling. We calculated the NTC for the different strategies defined as the number of patients that were counselled by a clinical geneticist divide by the number of PGVs detected.

## Results

### Overall germline findings in the WIDE cohort

Paired germline and tumour genome sequencing analysis was performed for 937 patients included in the WIDE cohort. Based on an unbiased, pseudonymized screening of 49 established cancer predisposition genes (CPGs; see Supplemental Table 1), 2309 tumour variants, consisting of 2179 somatic and 130 germline variants were reported by the bioinformatics pipeline on the OncoAct genome sequencing report (Figure 1, Supplemental Table 2,3). After revision 1509 (likely) pathogenic somatic and 109 (likely) pathogenic germline variants (PGVs) remained. The 109 PGVs were in 25 different genes, in 106 unique patients (11.6% of all patients). 25 monoallelic variants in recessively inherited predisposition genes *MUTYH* (n=21) and *NTHL1* (n=4) were excluded as only biallelic or compound heterozygous germline variants are considered having associated hereditary risks. In total, 84 PGVs (including clinically irrelevant PGVs, i.e. in tumour types not matching the per gene associated tumour type) in 23 different genes were present in 82 unique patients (9% of all patients) across 17 different primary tumour locations (Figure 2, Supplemental Table2).

**Figure 1.**
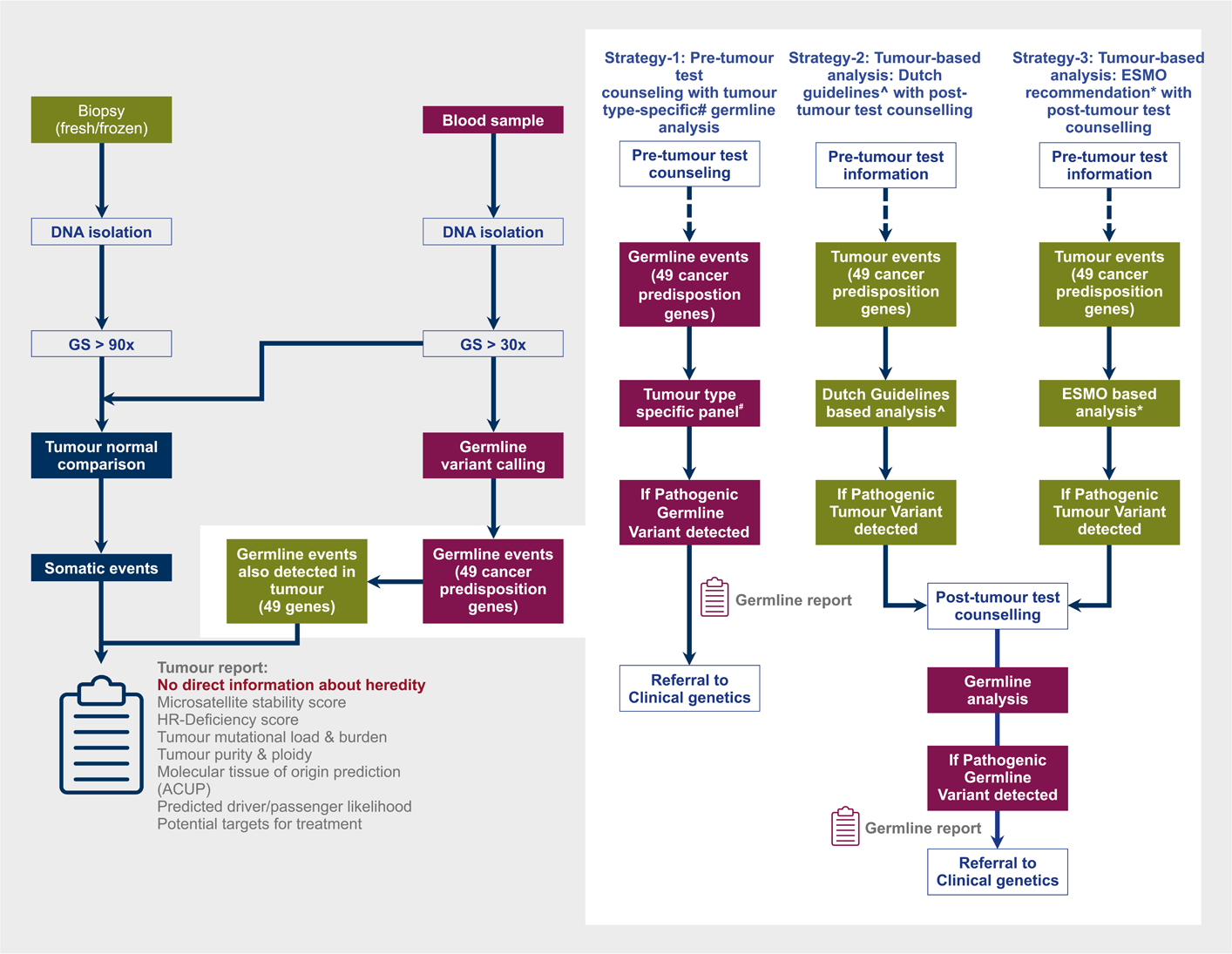
Overview of our genome sequencing (GS) pipeline with three strategies of integrated germline analysis. In general variants that are present in the germline and tumour are excluded from the genome sequencing report, except for variants in 49 established cancer predisposition genes. Variants in these genes based on reference genome version GRCh37/hg19 that are present in both germline and tumour are included on the tumour report as tumour variants (gene reference transcripts as indicated in Supplemental Table 1). These are analysed according to the three different strategies as indicated. #Tumour type-specific gene panel – see supplemental table 1. *ESMO PMWG germline recommendations – see reference [^3, 4^] and supplemental table 1. ^Adjusted Dutch guideline – see reference [^36^] and supplemental table 1.

**Figure 2.**
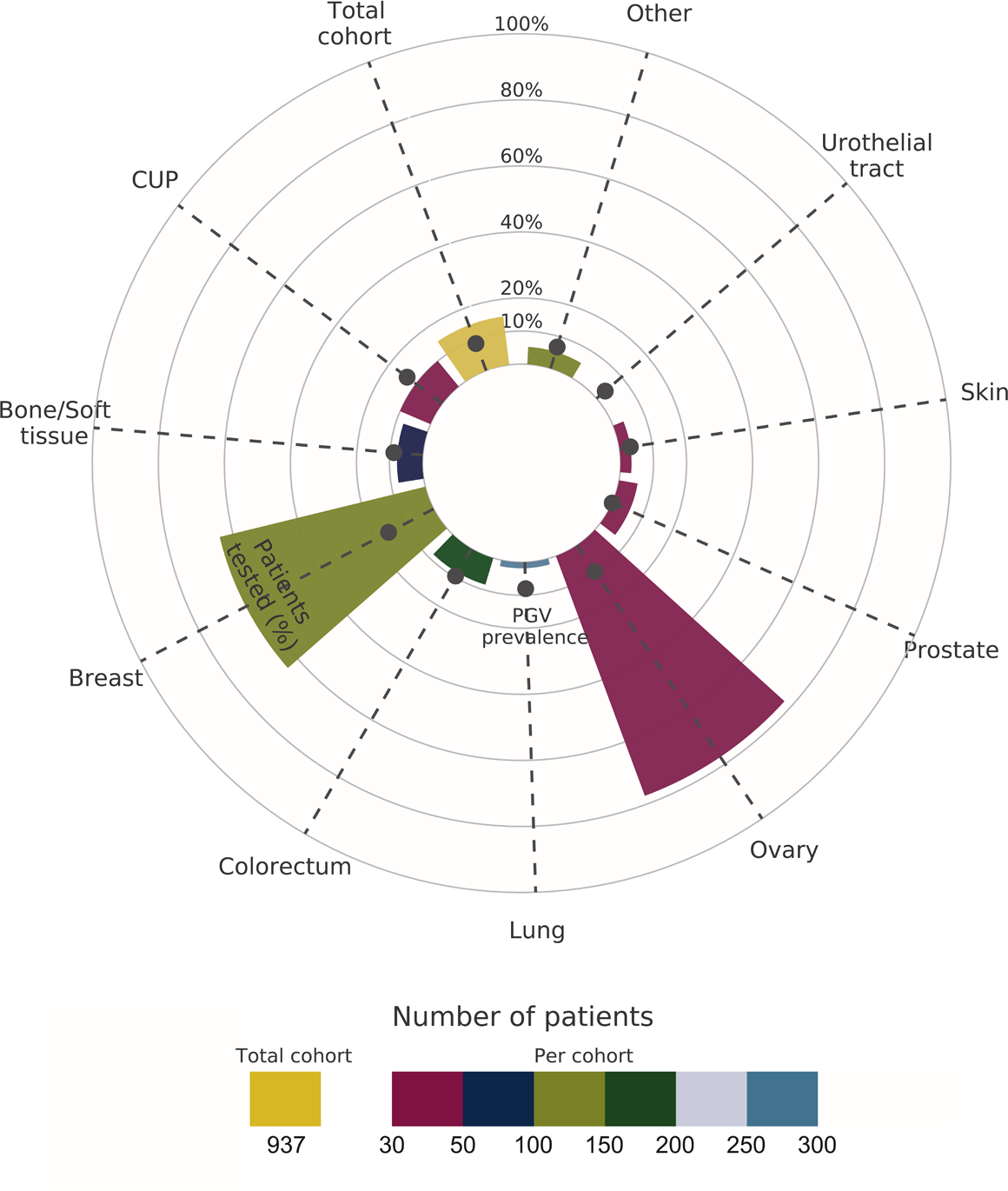
Prevalence of pathogenic germline variants per primary tumour location. Overview of percentage of patients with standard-of-care genetic testing prior to the genome sequencing (bars) and prevalence of pathogenic germline variants (dots) detected with the genome sequencing per primary tumour location/type. Locations with fewer than 30 patients are grouped in “other”. Bars are highlighted based on the number of patients. Data based on reference genome version GRCh37/hg19 and gene reference transcripts as indicated in Supplemental Table 1.

Prior to genome sequencing analysis, 138 patients had already received standard of care genetic (germline) testing as part of clinical guidelines^19^, familial cascade testing, or a potential germline finding in a tumour NGS test. Genetic testing was primarily performed in patients with ovarian cancer and breast cancer in 77.4% (24 of 31) and 64.1% (75 of 117) of the patients, respectively. In other common tumour types, like (non-small cell) lung cancer (5 of 274; 1,8%) and colorectal cancer (14 of 167; 8,4%), genetic testing was less often performed (Figure 2, Supplemental Table2). 28 PGVs were previously detected with genetic testing in eight different tumour types and detected using genome sequencing. Additionally, two PGVs were previously unrecognized (not covered in the gene panel at the time), though detected with genome sequencing. The prevalence of PGVs in this cohort that did receive genetic testing is therefore 21.7% (28+2/138).

Taken together, in the total WIDE cohort, 84 PGVs were present in 82 unique patients (9% of all WIDE patients) across 17 different primary tumour locations (Figure 2, Supplemental Table2).

### Strategy-1: Pre-tumour test counselling with direct tumour type-specific germline analysis

In strategy 1, all 937 patients need to be counselled about possible germline findings before the genome sequencing test. PGVs will be reported after simultaneous analysis of the germline based on the tumour type-specific gene panel (Figure1, Supplemental Table 1). This panel closely resembles the standard of care germline testing gene panels used for the specific tumour type(s) as indicated (Supplemental Table 1) ^19–22^. 50 PGVs in 50 patients would be reported resulting in a yield of 61% compared to baseline (50 of 82 unique patients – Figure 3, 4, Table 1), an NTC of 18.7 (937 total patient counselled / 50 patients with PGV – Figure 3, 4, Table 1), and a prevalence of 5.3% of detected PGVs in the whole WIDE study (50/937 – Table 1, Supplemental Figure 3,4). 25 out of 50 PGVs detected using strategy1 were found in 25 patients without prior genetic testing, including in patients that do not meet current genetic testing criteria.

**Figure 3.**
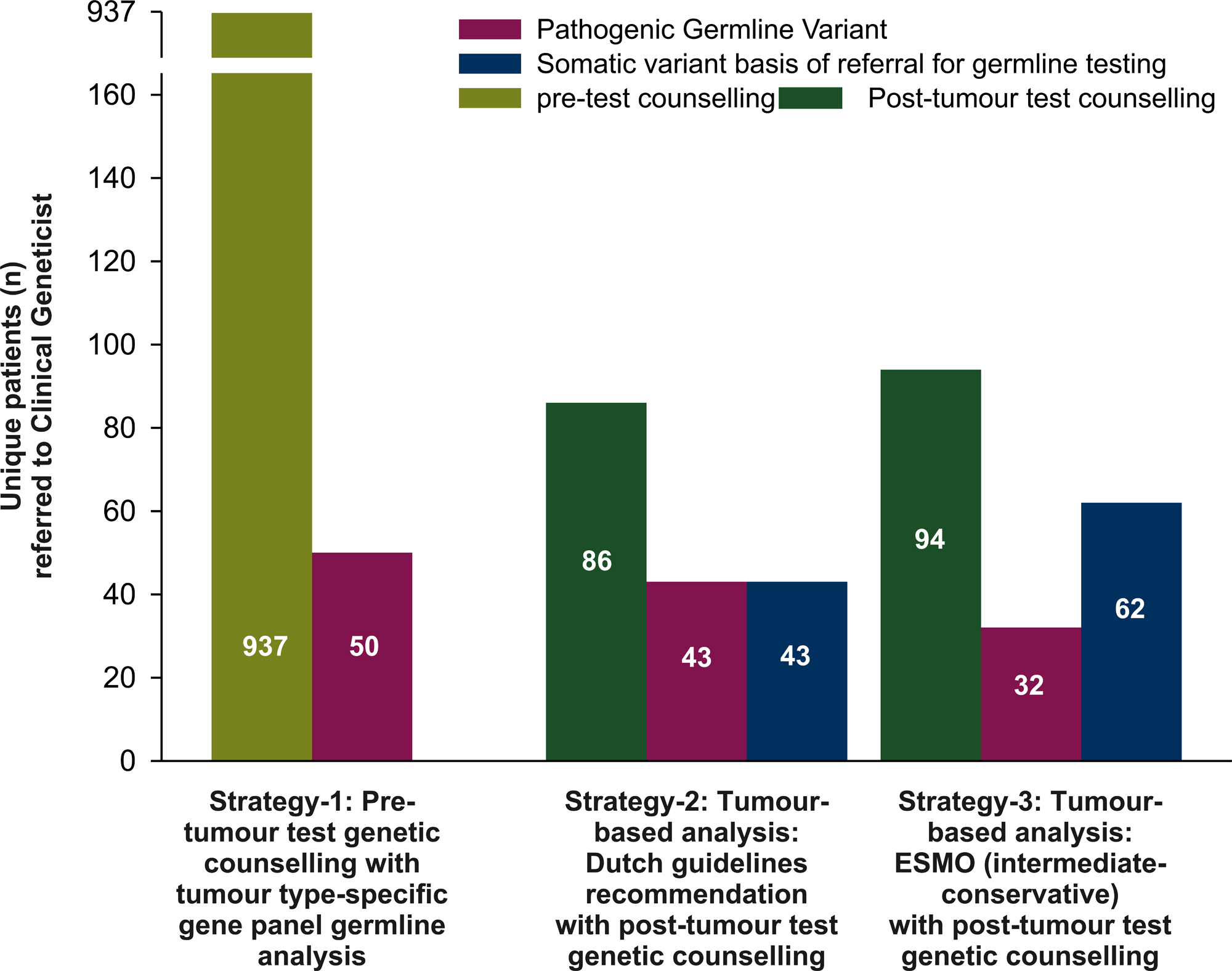
Referral to clinical geneticist following the three different strategies. Data based on reference genome version GRCh37/hg19 and gene reference transcripts as indicated in Supplemental Table 1.

**Table 1.** Pathogenic variants detected, number-to-counsel and PGV yield.

|  | Pathogenic Germline variants (n) | Pathogenic Somatic variants (n) | Total (n) pathogenic variants | Patients with PGV (n) | Patients with somatic variants (n) | Patients counselled (n) | Prevalence (%) | NTC | Yield (%) | Yield/NTC |
| --- | --- | --- | --- | --- | --- | --- | --- | --- | --- | --- |
| Baseline | 84 | 0 | 84 | 82 |  | 937 | 9.0 | 11.4 | 100 | 8.8 |
| Strategy-1 – direct germline analysis with Tumour specific gene panel | 50 | 0 | 50 | 50 |  | 937 | 5.3 | 18.7 | 61 | 3.3 |
| Strategy-2 Adjusted Dutch guidelines | 43 | 63 | 106 | 43 | 43 | 86 | 4.6 | 2.0 | 52 | 26.2 |
| Strategy-3 ESMO 2022 Intermediate-conservative | 32 | 92 | 124 | 32 | 62 | 94 | 3.4 | 2.9 | 39 | 13.3 |
| ESMO 2018/19 | 32 | 101 | 133 | 32 | 69 | 101 | 3.4 | 3.2 | 39 | 12.4 |
| ESMO 2022 permissive | 62 | 202 | 264 | 61 | 134 | 195 | 6.6 | 3.2 | 74 | 23.3 |
| ESMO 2022 Intermediate-permissive | 37 | 115 | 151 | 37 | 79 | 116 | 3.9 | 3.1 | 45 | 14.4 |
| ESMO 2022 Conservative | 20 | 70 | 90 | 20 | 46 | 66 | 2.1 | 3.3 | 24 | 7.4 |
| Hybrid form germline analysis of 6 core genes + |  |  |  |  |  |  |  |  |  |  |
| Adjusted Dutch guideline for other genes | 48 | 18 | 66 | 48 | 16 | 64 | 5.1 | 1.3 | 59 | 43.9 |
| ESMO 2022 Intermediate-permissive for other genes | 37 | 42 | 79 | 37 | 32 | 69 | 3.9 | 1.9 | 45 | 24.2 |

NTC (number-needed-to-counsel; Patients counselled (n)/patients with PGVs (n).

Compared to the baseline, 34 PGVs in 32 patients would remain unreported, since these were considered irrelevant per tumour type-specific gene panel (Figure 3, 4, Table 1). The majority of unreported PGVs were in tumour type-unspecific intermediate-risk genes (20 total; 14x *CHEK2* and 6x *ATM*). Furthermore, 14 PGVs were in tumour type-unspecific high-risk genes, including PGVs in *BRIP1*; (n=3) *BARD1 (n=2), TP53 (n=2)* and *NF1* (n=2; *see* Figure 3, 4, Supplemental Table 2). Interestingly, three of 14 PGVs in the tumour type-unspecific high-risk genes had somatic LOH (1x *BAP1*, 1x *TP53* and 1x *NF1*), indicating that those might have played a role in the tumour development.

### Strategy-2: Referral for post-tumour test genetic counselling and testing based on presence of a pathogenic tumour variant per adjusted Dutch guidelines

The second strategy is a tumour-based analysis with subsequent referral for post-tumour test genetic counselling on germline analysis and testing based on pathogenic tumour variants (PTV) present in the genome sequencing report. This strategy was guided by adjusted Dutch guidelines (Figure 1, Supplemental Table 1)^36^. 106 PTVs were present in 86 unique patients. All 86 patients would be referred to the clinical genetics department for counselling and after consent germline-testing would be performed (Figure 3, 4, Table 1). After germline-testing, 43 PTVs appeared to be clinically relevant PGVs in 43 patients would be detected resulting in a 52% yield compared to baseline (43 of 82 unique patients – Figure 3, 4, Table 1), an NTC of 2 (86 patients counselled / 43 patients with PGV) and a prevalence of detected PGVs of 4.6% in the overall WIDE cohort (43/937 – supplemental Figure 1,2, Table 1). This also means that 63 PTVs in 43 unique patients were most likely of somatic origin and 43 patients were unnecessarily referred for counselling. The majority of somatic variants were in the homologous recombination repair (HRR; 28 variants in total; n=21 in *BRCA2* and n=7 in *BRCA1*) and mismatch repair (MMR; 18 variants in total, including n=10 in *MSH6*) genes (Figure 4).

**Figure 4.**
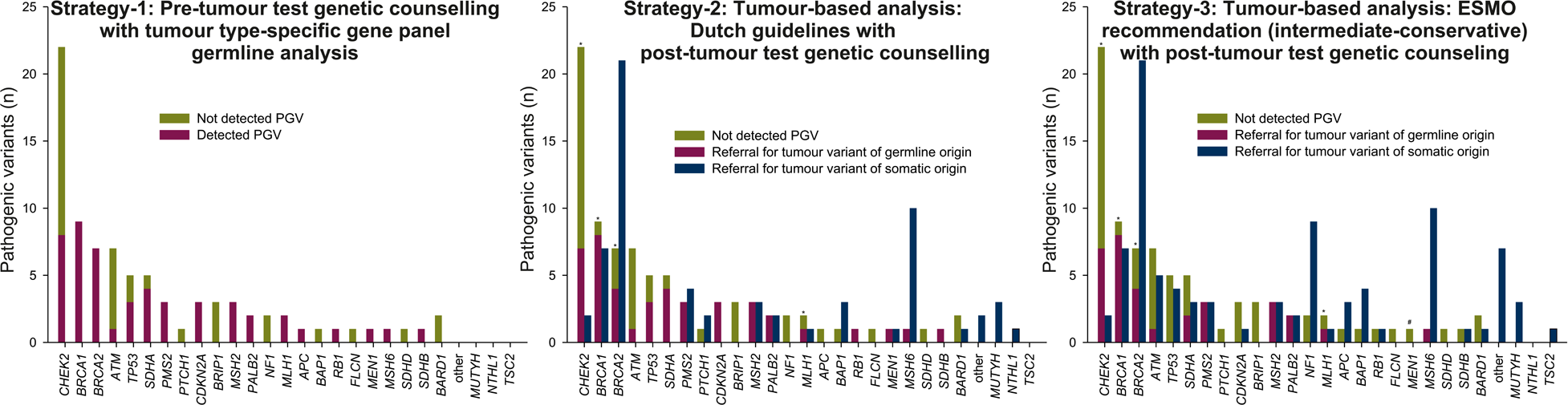
Pathogenic variants (un)detected for the three different strategies on gene level. * 3x *BRCA2*, 1x *BRCA1*, 1x *MLH1* and 1x *CHEK2* are somatically lost in the tumour and would have been reported if they were present on the tumour report per Adjusted Dutch guidelines and ESMO PMWG recommendation. # ESMO PMWG recommendation has no recommendation for *MEN1*. Data based on reference genome version GRCh37/hg19 and gene reference transcripts as indicated in Supplemental Table 1.

### Strategy-3: Referral for post-tumour test genetic counselling and testing based on presence of a pathogenic tumour variant per ESMO PMWG

The third strategy is a tumour-based analysis with subsequent referral for post-tumour test genetic counselling on germline testing and testing based on pathogenic tumour variants (PTV). This strategy was guided by the ESMO PMWG recommendation, using both the former and updated (four-tiered) recommendations (Figure 1, Supplemental Table 1)^3, 4^. Here we show data from the intermediate-conservative stringent updated ESMO PMWG recommendation because this is recommended by the ESMO PMWG for European countries (for the other ESMO PMWG tiers see Table 1). 94 patients with 124 PTVs would be referred to the clinical genetics department for counselling and after consent germline-testing would be performed. After germline-testing, 32 PGVs in 32 unique patients would be confirmed in the germline with a 39% yield compared to baseline (32 of 82 patients – Figure 3, 4, Table 1), an NTC of 2.9 (94 patients counselled / 32 patients with PGV) and an overall prevalence of pathogenic germline variants of 3.4% in the overall WIDE cohort (32/937 – supplemental Figure 3,4). Notably, 92 PTVs of the 124 PTVs in 62 unique patients were most likely of somatic origin. The majority of pathogenic variants of somatic origin were in the HRR (28 variants in total; n=21 in *BRCA2* and n=7 *BRCA1*), MMR (17 variants in total, including n=10 in *MSH6*), *NF1* (n=9) and *TP53* (n=4) genes (Figure 4).

### Difference between the three strategies

In strategy-2 (adjusted Dutch guidelines), compared to strategy-1 (pre-tumour test counselling with tumour type-specific directly germline analysis) PGVs in seven patients were missed, including six PGVs that were somatically lost in the tumour and therefore not reported in the somatic tumour report (see below). (Supplemental table 4). Additionally, one PGV in the high-risk gene *APC* was not referred for a patient with Colorectal cancer at age 49, since *APC* is only considered when the patient and/or family members have polyposis (Supplemental table 5).

In strategy-3 (ESMO-PMWG intermediate-conservative tier), compared to strategy-1, PGVs in 18 patients were missed, again including the same six PGVs that were somatically lost in the tumour. Additionally, 12 patients with PGVs in high-risk genes were not referred, including PGVs in *TP53* (n=3), *RB1 (*n=1) and *APC* (n=1) since the patients did not match ESMO-PMWG’s age requirement (any tier requires matching tumour type and age < 30 year – see Supplemental table 5). Furthermore, one PGV in *MEN1* was not referred since ESMO-PMWG has no recommendation for *MEN1*.

Interestingly, six PGVs were missed with strategy 2 and 3 because the allele harbouring the PGV was somatically lost in the patients tumour (3x *BRCA2*, 1x *BRCA1*, 1x *MLH1* and 1x *CHEK2*). These variants were detected in the germline-based strategy-1 because in this strategy we could report hereditary predisposition directly based on germline analysis (blood). They were not detected with the other two strategies however, because our genome sequencing pipeline does not report PGVs that are lost in the tumour. The reasoning behind this is that for these variants the role in the oncogenesis is uncertain.

4 of the 5 patients with a *BRCA1/2* or *MLH1* variant with an associated HRD or MSI signature, all exhibited a homologous repair or microsatellite proficient profile (Supplemental table 4). This lack of the associated signature might indicate that the PGV did not play a role in the oncogenesis of the tumour. However, 1 of the patients with a lost pathogenic *BRCA2* germline variant did show an HRD profile. This patient however was previously treated with a PARP inhibitor and the loss of the *BRCA2* germline variant in this tumour fits with the scenario where the mutated allele was selected against due to therapeutic pressure^46^. For the 6^th^ patient with a *CHEK2* PGV there are currently no associated tumour signatures. Importantly, in 5 out 6 patients these missed PGVs were clinically relevant per tumour type or family history in the context of hereditary predisposition (Supplemental table 4).

## Discussion

In this study, we compared the PGV yield and associated number-needed-to counsel in 937 tumours analysed by genome sequencing for three different strategies. A pre-tumour test counselling strategy directly analysing the germline (strategy-1) and two post-tumour test counselling strategies (based on the adjusted Dutch guidelines [strategy-2]^36^ and the ESMO PMWG recommendation [strategy-3]^3, 4^). Results show the highest yield of 61% for strategy-1 vs. 52% and 39%, for strategy-2 and strategy-3 respectively.

In comparison to strategy-1, a considerable proportion of PGVs in strategy-2 (6 out of 7) and strategy-3 (6 out of 18) were not detected due to somatic loss of 6 PGVs in the associated tumours. This is an important disadvantage of strategies 2 and 3. In the context of molecular tumour analysis it seems valid not to report these variants because 5 out of 6 might not play a role in the tumour oncogenesis of the tested tumour and are therefore probably not relevant to the tumour treatment. Patients could even be over treated based on a germline variant not present in the tumour. However in the context of screening strategies that aim to identify all hereditary predisposition by sequencing the tumour (e.g. Tumour First ^47^) this poses a challenge because a relevant hereditary predisposition could be missed. We therefore also looked at the tumour types and available family history in our database of these six patients to see if the ‘missed’ PGVs were clinically relevant. Indeed in 5 of the six patients the PGVs was relevant for familial cascade testing because either the PGV was not detected in a relevant tumour (e.g. *BRCA2* in a breast carcinoma) or the family history showed matching tumour types that warrant further follow up (Supplemental table 4).

Theoretically, the lost PGVs could easily be added to the tumour report. This would increase the diagnostic yield to 60 and 46% for strategy 2 and 3 respectively which makes the yield of strategy-2 almost comparable to strategy-1 (61% yield), since only 1 PGV in *APC* is now missed. However, one should be aware that by altering the pipeline this way, variants that appear in the tumour report can be germline heterozygotes without a role in tumour oncogenesis and/or treatment, which may be confusing for the interpretation. Therefore, careful consideration is necessary before including the lost in tumour PGVs on the report. It is important to emphasize that regular NGS panels also report these lost PGVs based on detection in normal tissue surrounding the tumour tissue. So, depending on the tumour cell percentage these NGS panels will or will not detect these PGV’s in the surrounding normal tissue.

The highest yield of the pre-tumour test counselling approach requires adequate genetic counselling of all patients prior to paired tumour-germline sequencing. As a result, the number-needed-to counsel (NTC=18.7) is high, especially compared to both post-tumour test strategies that have an NTC of 2 (adjusted Dutch guidelines) and 2.9 (ESMO PMWG) respectively (Table 1).

When comparing post-tumour test counselling strategy-2 and strategy-3, the number of patients referred for genetic counselling and the NTC was higher for the permissive, intermediate-permissive, intermediate-conservative and “old” ESMO PWMG recommendations (strategy-3) as compared to adjusted Dutch guidelines (strategy-2; see Tabel1). Simultaneously, less PGVs were detected using the ESMO PWMG recommendations as compared to adjusted Dutch guidelines (Table 1). Both the former and recently updated recommendations from the ESMO PWMG are largely constituted around the per gene germline conversion rate (GCR), i.e., the rate of which a tumour variant in a gene of interest can be attributed to an underlying pathogenic germline variant. For genes with very common somatic variants, like *TP53* or *APC*, the per gene GCR is intrinsically low. As a result, follow-up genetic testing is only recommended in specific cases (on-tumour setting, age < 30 year), to prevent large-scale genetic testing in low a priori risk populations. Thus, the per gene GCR is used as surrogate for clinical information and family history. In contrast, the adjusted Dutch guidelines uses clinical/familial criteria to pin-point specific cases to follow-up. The difference in approach is especially noticeable for *TP53.* With the adjusted Dutch guidelines, three patients with a PGV in *TP53* would have been detected, while no patients would have been referred based on a somatic *TP53* variant. On the other hand, using the ESMO PWMG recommendation four patients with a somatic variant in *TP53* would have been referred and no patients with a PGV in *TP53* were detected (Figure 4).

Current screening strategies based on genetic testing criteria can be complex^18^. Furthermore PGVs are detected in patients without clinical indication as shown in this study and by many other studies^6–18^. Evidence is cumulating that a tumour/germline based approach is necessary to increase PGV yields and thus there is an increasing need for a more universal approach^18, 48, 49^. However, directly analysing the germline requires genetic counselling and informed consent of the patient. It would impose too heavy of a burden on genetic counsellors to offer pre-test counselling too all these patients. Currently, mainstreaming of genetic care, in which pretest counselling by nongenetic health care professionals is integrated into routine care, is studied by multiple groups and seems feasible and resource efficient^50–58^. Mainstreaming has many advantages, all patients (that consented) can be included and genetic counsellors only get patients referred with a proven PGV. Also, all data relevant for diagnostic purposes and/or treatment strategy will be available at a single time point. A disadvantage of mainstreaming is that it requires sufficient knowledge and skills on genetic counseling for all requesting professionals and thus education and training is essential^59^. Moreover, it is important to note that pretest counseling of a limited set of genes, i.e. a breast and ovarian cancer gene panel, requires a different level of genetic counseling expertise compared to counseling a broad oncogenetic panel in which many different types of cancer predisposition genes may be identified with different associated levels of cancer risks and variable screening (im)possibilities. Importantly, compared to the US, Europe is more restrictive and the proposed language suggested by the ACMG^60^ might not suffice for certain genes (e.g. for syndrome specific genes such as *TP53*), albeit the remark that the patients can consult a genetic counselor for more information.

In the Netherlands Cancer Institute, a comprehensive cancer centre, we have implemented genome sequencing in routine clinical cancer care since January 2021. For the reasons mentioned above, we therefore, have explored multiple strategies of integrated germline analysis to learn which strategy fits best in our routine clinical care. Genome sequencing results are discussed in a weekly dedicated molecular tumour board, consisting of at least a Pathologist, Clinical Scientist in Molecular Pathology, Clinical Geneticist, Clinical Laboratory Geneticist and an Oncologist. Referral for genetic counselling is communicated by direct contact of the clinical geneticist with the treating physician and by an additional remark in the routine pathology report (Figure 5). We now use a post-tumour test counselling approach using the adopted adjusted Dutch guidelines for referring patients to the clinical geneticist, since it has the best PGV yield per NTC, especially because clinical information and family history is readily available in our hospital during evaluation of the genome sequencing data.

**Figure 5.**
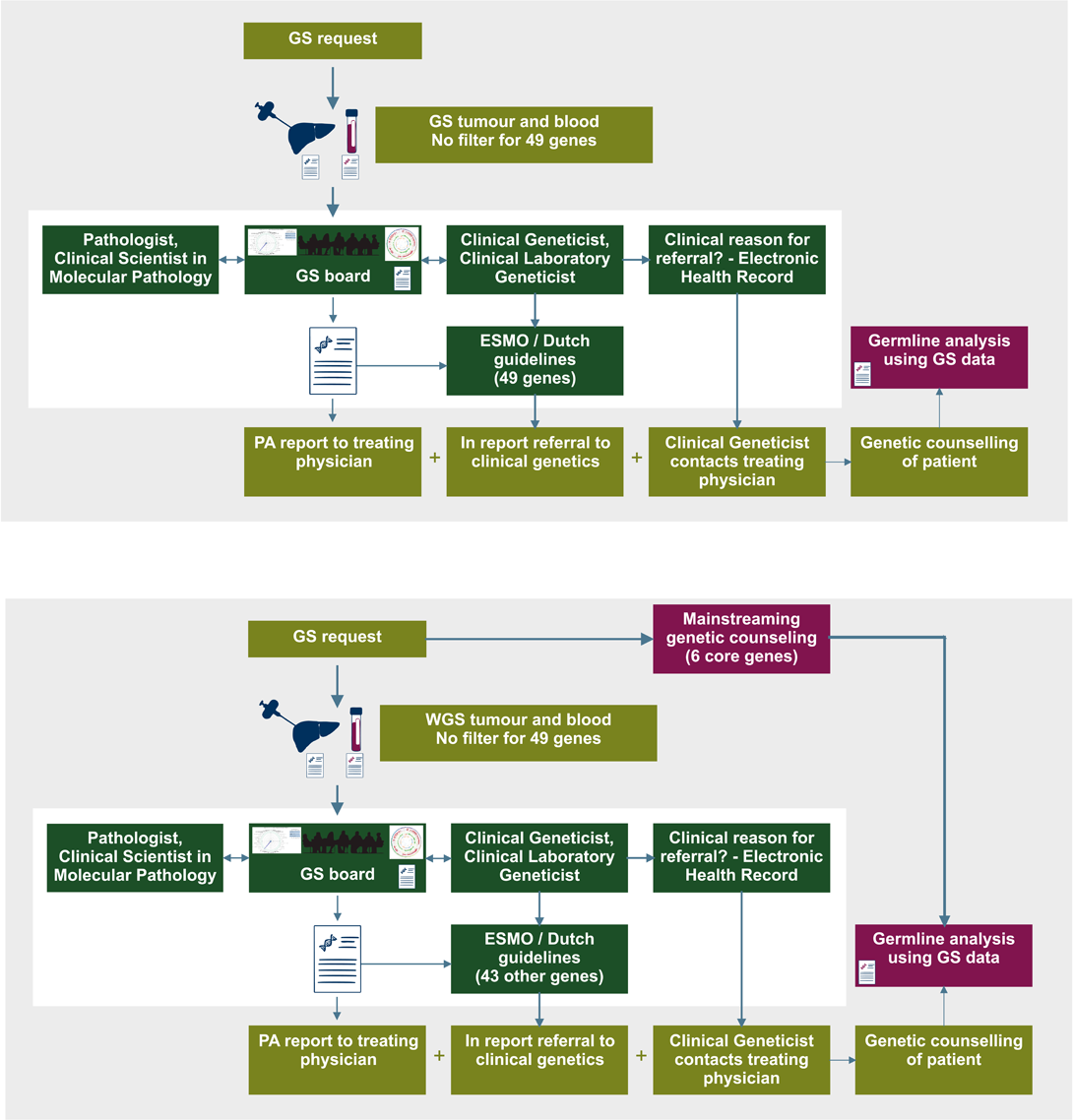
Overview of our dedicated molecular tumour board and proposed changes. Genome sequencing (GS) results are discussed in a weekly dedicated molecular tumour board, consisting of at least a pathologist, clinical scientist in molecular pathology, clinical geneticist, clinical laboratory geneticist and an oncologist. Referral for genetic counselling is communicated by direct contact of the clinical geneticist with the treating physician and by an additional remark in the routine pathology report. **A**, current practice in which we use a post-tumour test counselling approach using the adopted Dutch guidelines for referring patients to the clinical geneticist. **B**, proposed hybrid form with pre-test mainstreaming for *BRCA1/2, PALB2, MLH1* and *MSH2/6* genes combined with post-test counselling for other remaining cancer predisposition genes.

Our current multidisciplinary approach using the adjusted Dutch guidelines has multiple advantages. First, compared to strategy 1, where the direct reporting is integrated in the reporting pipeline per tumour type, we can instantly switch gears e.g. when the genome sequencing data points to a not expected tumour type or in the context of a cancer of unknown primary (CUP) patient^61^.

Further, with the availability of additional clinical information (e.g. history of other primary tumour and/or tumour in other family members) the clinically relevance of specific genes for germline follow-up and therapeutic decisions might change.

For instance, a male with an abdominal metastasized CUP, a PGV in *CHEK2* became clinically relevant per adjusted Dutch guideline for germline follow-up, since his sister was diagnosed with breast cancer. In addition to the referral to the clinical genetics department, he became eligible for experimental PARPi treatment based on the complete loss of *CHEK2* on his tumour report. Likewise, for a male patient with lung cancer, detection of a PGV in *ATM* with increased risk for breast cancer in females did not have any clinical relevance for germline follow-up, as this patient did not have any living female relatives. However, based on the complete loss of *ATM* in the genome sequencing tumour report, this patient became eligible for experimental PARPi treatment. Thus, the diagnostic yield of the adjusted Dutch guidelines could further increase when familial data is taken into account.

We noticed, in our simulated WIDE study data, that the majority of pathogenic somatic and germline variants were within six cancer predisposition genes (*BRCA1*, *BRCA2*, *PALB2*, *MLH1*, *MSH2* and *MSH6;* see Figure 4) that are considered relevant to refer for any tumour type using both ESMO PWMG recommendation and adjusted Dutch guidelines. Regarding the future workload for genetic counsellors and the already long waiting lists for patients and possibly associated psychological burden e.g. waiting for test results, we hypothesized how we could further reduce unnecessary referrals while detecting most of the relevant PGVs and offer sufficient pre-test genetic counselling. We therefore propose a hybrid form where directly analysing the germline for the above six genes in combination with one of the tumour-based strategies for the remaining cancer predisposition genes (Figure 5B) would result in a higher PGV yield and less unnecessary referrals for a somatic variant. For instance, this hybrid approach combined with the adjusted Dutch guidelines, would have led to a yield of 59% (compared to 52% for only adjusted Dutch guidelines) and 27 patients with a somatic variant would not be unnecessary referred for germline testing (Table 1; Figure 5). Using this hybrid approach patients need to be specifically pre-tumour test informed by mainstreaming for only these six genes that can be performed by the treating physician or other trained staff supported with standardized educational material. The proposed language as suggested by the ACMG^60^ is perfectly suited for this.

In summary, with this study we benchmarked a pre-tumour test counselling and two post-tumour test counselling strategies to incorporate germline analysis into tumour diagnostic sequencing. All three strategies had a comparable detection rate of clinically relevant pathogenic germline variants, but with different number-needed-to counsel. With clinical information and family history available, the adjusted Dutch guidelines has the best PGV yield per NTC. A combination of pre-tumour test mainstreaming and post-tumour test counselling may maximize the clinically relevant PGV yield of true germline variants and minimize unnecessary referrals. Further studies are necessary regarding ethical aspects, cost efficiency and patient and professional experiences.

## Ethics Declaration

The WIDE study was performed in concordance with the Declaration of Helsinki, Dutch law, and Good Clinical Practice after approval by the Medical Ethical Committee of the Netherlands Cancer Institute (NKI) (NL68609.031.18). All included patients provided written consent for study participation.

## Author contributions statement

RK, NG, TW, LK and KM were responsible for the conceptualization of the study design. EC, EV, GM and KM were involved in funding acquisition. RK, LS, NG, DB, MM, KS, ER, FH, PR, MB, LB, JB, MR, TW, LK and KM for data collection and/or were responsible for part of the data analysis. RK, LS, NG and KM were responsible for drafting of the article. The underlying data reported in the article has been accessed and verified by multiple authors (RK, LS, and KM). All authors have read, revised, and approved the article.

## Data availability statement

The researchers are willing to share with external qualified researchers access to patient-level data and supporting clinical documents. These requests are reviewed and approved by an independent review committee on the basis of scientific merit. All data provided are anonymized to respect the privacy of patients who have participated in the study, in line with applicable laws and regulations. Data can be requested via https://www.hartwigmedicalfoundation.nl/applying-for-data/ or.

## Disclosure

KM reports research grants from AstraZeneca and speakers’ fees from MSD, Roche, AstraZeneca. and Benecke. KM received consultancy fees from Pfizer, BMS, Roche, MSD, Abbvie, AstraZeneca, Diaceutics, Lilly, Bayer, Boehringer Ingelheim and nonfinancial support from Roche, Takeda, Pfizer, PGDx and DELFi. EC reports consultancy fees and support for attending meetings and traveling from Illumina. EV is member of the supervisory board of Hartwig Medical Foundation. GM is cofounder and a board member (CSO) of CRCbioscreen BV, he has a research collaboration with CZ Health Insurances (cash matching to ZonMW grant) and he has research collaborations with Exact Sciences, Sysmex, Sentinel Ch. SpA, Personal Genome Diagnostics (PGDX), DELFi; these companies provide materials, equipment, and/or sample/genomic analyses. GM is an Advisory Board member of ‘Missie Tumor Onbekend.’ RK, LS, NG, DB, MM, KS, ER, FH, PR, MB, LB, JB, MR, TW, and LK report no conflicts of interest.

## Supplemental information

Supplemental Figure 1. Variants in 57 genes that have diagnostic and/or therapeutic relevance that are present in both germline and tumour are included on the genome sequencing report.

Supplemental Figure 2. 20 DNAs from blood with a complex and/or difficult to detect germline finding from SOC diagnostics were reanalysed using genome sequencing.

Supplemental Figure 3. Prevalence of pathogenic germline variants per primary tumour location using the strategy as indicated.

Supplemental Figure 4. Prevalence of pathogenic germline and somatic variants per primary tumour location using the strategy as indicated.

Supplemental Table 1. Gene list and information.

Supplemental Table 2. All germline findings

Supplemental Table 3. All findings (somatic and germline)

Supplemental Table 4. Germline findings that are somatically lost

Supplemental Table 5. Differentially referred germline findings

## Data Availability

All data produced in the present work are contained in the manuscript

https://www.hartwigmedicalfoundation.nl/en/data/data-acces-request/

