## Supplemental Figures for "Impact of genetic counselling strategy on diagnostic yield and workload for genome sequencing-based tumour diagnostics"

**Supplemental Figure 1.**
Variants in 57 genes that have diagnostic and/or therapeutic relevance that are present in both germline and tumour are included on the OncoAct GS report as variants present in the tumour, although their germline status is not explicated annotated. 49 are established cancer predisposition genes (see Supplemental table 1). Data based on reference genome version GRCh37/hg19 and gene reference transcripts as indicated in Supplemental Table 1.

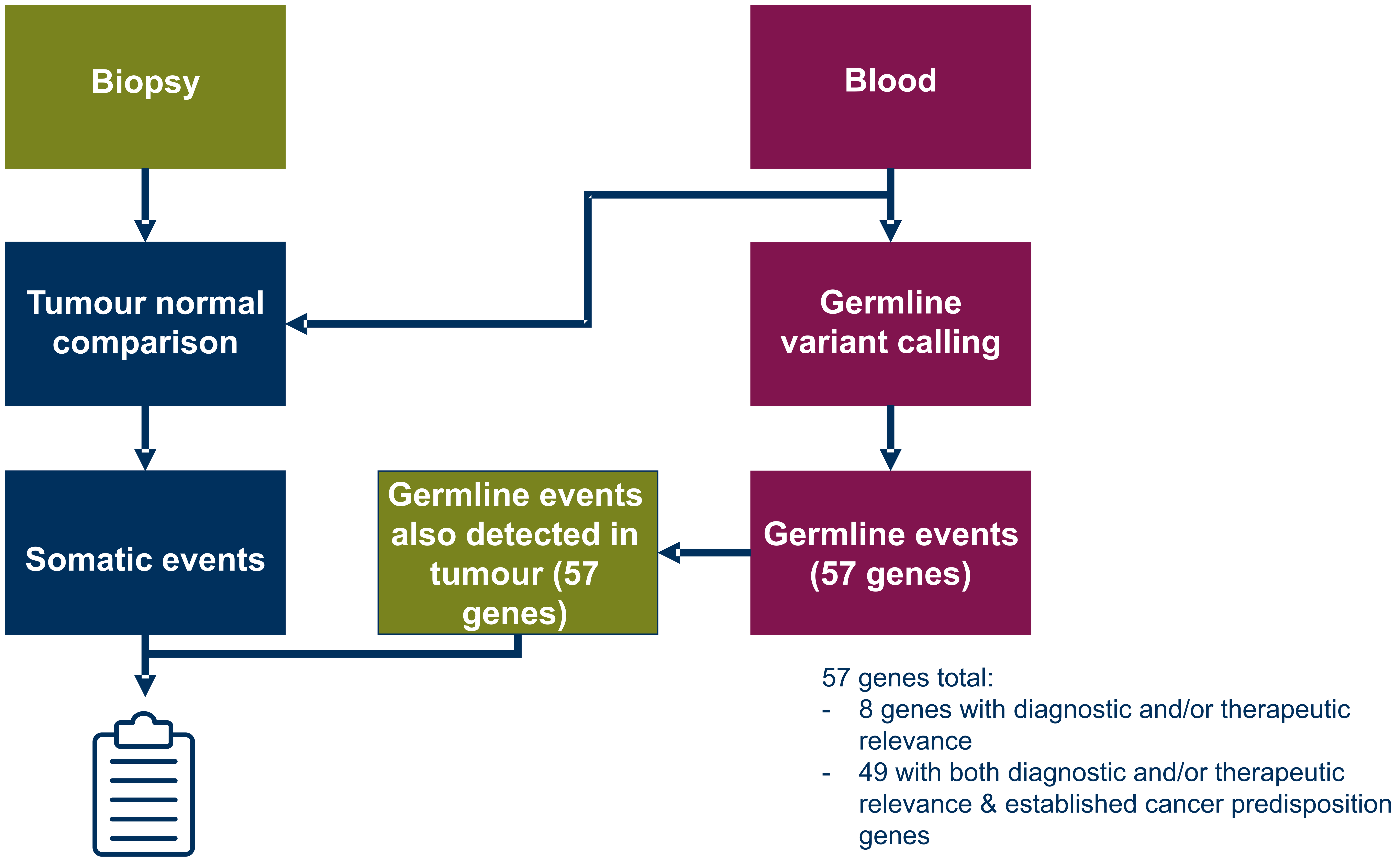

**Supplemental Figure 2.**
20 DNAs from blood with a complex and/or difficult to detect germline finding from SOC diagnostics were reanalysed using genome sequencing. All complex and/or difficult to detect variants were detected, although 2 deletions in PMS2 with low quality scores (table left side). In these 20 samples >99,5% (median and average) of bases within the gene panel is covered with > 20 reads (plot right side). Note that some of the sample had a major drop in coverage for certain genes, that corresponds with the gene deletion. Two examples of germline findings visualised using IGV are shown bottom left. 2x2 table used to calculate sensitivity and specificity is shown bottom left. Data based on reference genome version GRCh37/hg19 and gene reference transcripts as indicated in Supplemental Table 1.

| Sample ID | Germline variants (as determined by SOC) | Additional difficulties | Coverage (mean) genome sequencing | VCF Filter status |
|---|---|---|---|---|
| S21-0001 | *MSH2* :c.942+3A>T | repeat (A-stretch) | 32 | PASS |
| S21-0002 | *EPCAM* :c.(?_904-1)_*MSH2* :c.(211+1_212-1)del | | 34 | PASS |
| S21-0003 | *CDKN2A* :c.9_32dup | | 31 | PASS |
| S21-0004 | *MLH1* :c.(1039-?)_(1409+?)del | | 35 | PASS |
| S21-0005 | *MSH6* :c.3920_3927dup | | 33 | PASS |
| S21-0006 | *BRCA2* :c.9063_9078del | | 33 | PASS |
| S21-0007 | *BRCA1* :c.4186-1787_4358-1667dup | | 32 | PASS |
| S21-0008 | *PMS2* :c.2276-91_2445+790del | pseudo region | 30 | SGL breakend with low_qual |
| S21-0009 | *PALB2* :c.(?_-200)_(3113+1_3114-1)del | | 31 | PASS |
| S21-0010 | *BRCA2* :c.8633-207_8754+740dup | | 38 | PASS |
| S21-0011 | *PMS2* :c.(705+1_706-1)_(903+1_904-1)del | | 35 | SGL breakend with low_qual |
| S21-0012 | *PMS2* :c.2445+1G>A | pseudo region | 34 | PASS |
| S21-0013 | *MSH6* :c.3261dup | repeat (C-stretch) | 35 | PASS |
| S21-0014 | *MSH2* :c.(367-?)_(645+?)del | | 38 | PASS |
| S21-0015 | *BRCA1* :c.(?_-200)_(441+?)del | | 34 | PASS |
| S21-0016 | *BRCA2* :c.9190_9234delinsN[25] | | 33 | PASS |
| S21-0017 | *MSH6* :c.3647-3_3668dup | | 33 | PASS |
| S21-0018 | *PMS2* :c.(?_-87)_(*160_?)del | pseudo region | 40 | SGL breakend with PASS |
| S21-0019 | *BRCA1* :c.4186-1632_4357+2031del3835 | | 36 | PASS |
| S21-0020 | *BRCA1* :c.5333-36_5406+400del | | 43 | PASS |

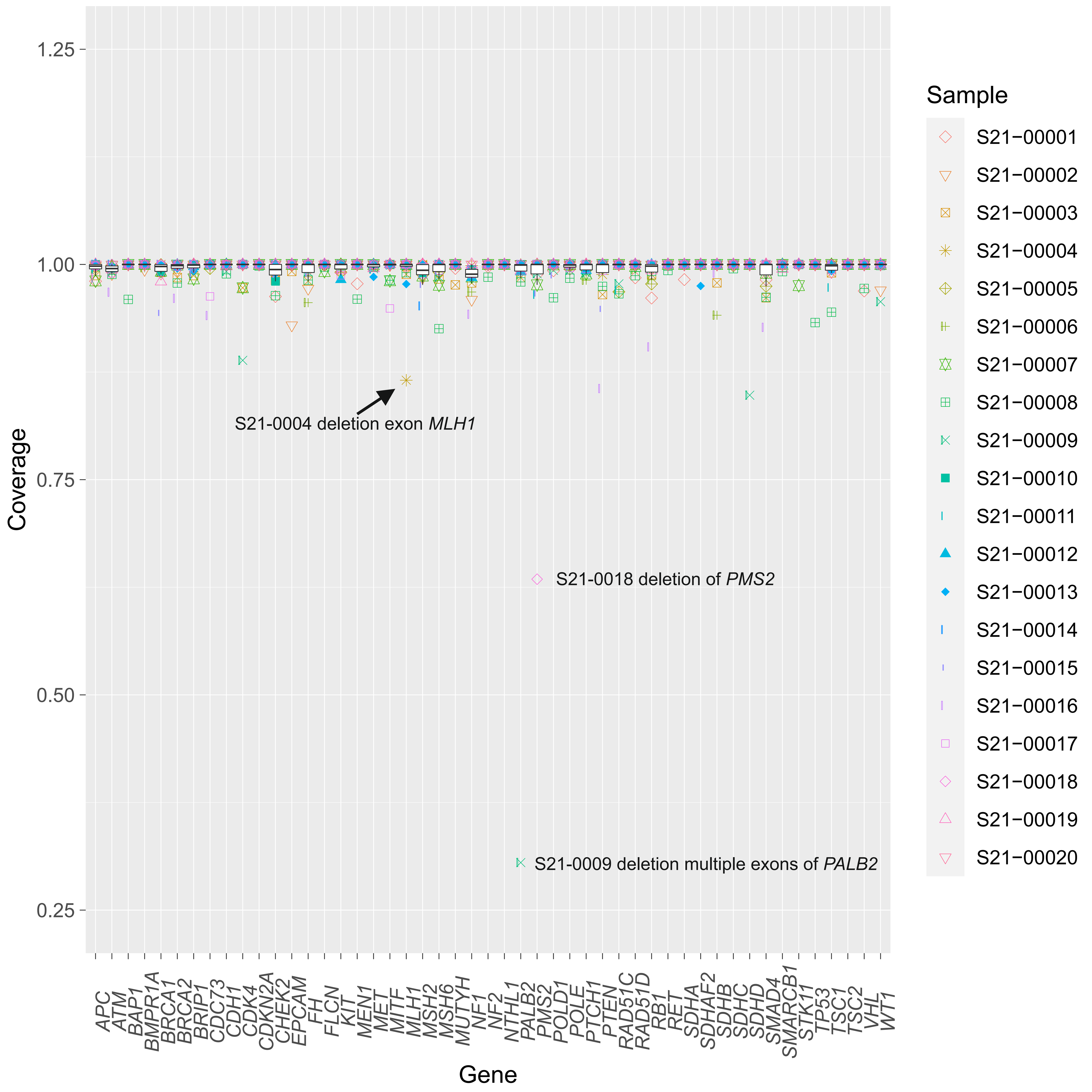


**S21-0001 – *MSH2* Splice variant in A-stretch**

***MSH2*:c.942+3A>T (VAF 46%)**

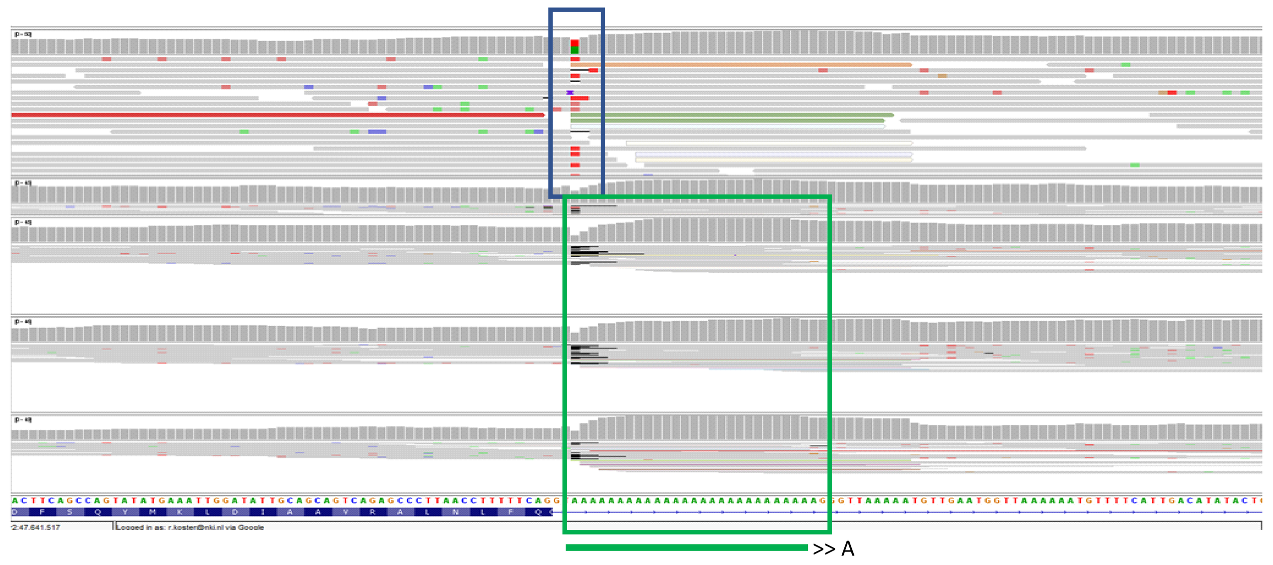

**S21-0008 - Del exon 14 *PMS2* (pseudo region)**

***PMS2*:c.2276-91_2445+790del**

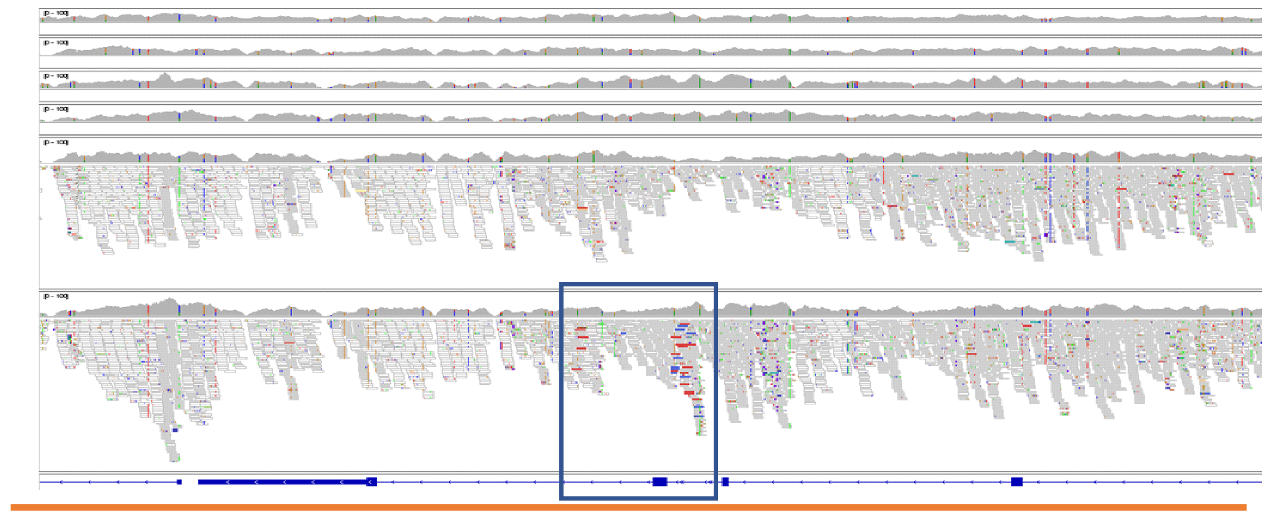


| | SOC - | SOC + | Totals |
|---|---|---|---|
| GS + | 0 | 87 | 87 |
| GS - | 137 | 0 | 137 |
| Totals | 137 | 87 | 224 |

**Supplemental Figure 3.**

Prevalence of pathogenic germline variants per primary tumour location using the strategy as indicated. Overview of percentage of patients with standard-of-care genetic testing prior to the GS (bars) and prevalence of pathogenic germline variants (dots) detected with the GS per primary tumour location/type using the strategy as indicated. Locations with fewer than 30 patients are grouped in “other”. Bars are highlighted based on the number of patients. Data based on reference genome version GRCh37/hg19 and gene reference transcripts as indicated in Supplemental Table 1.

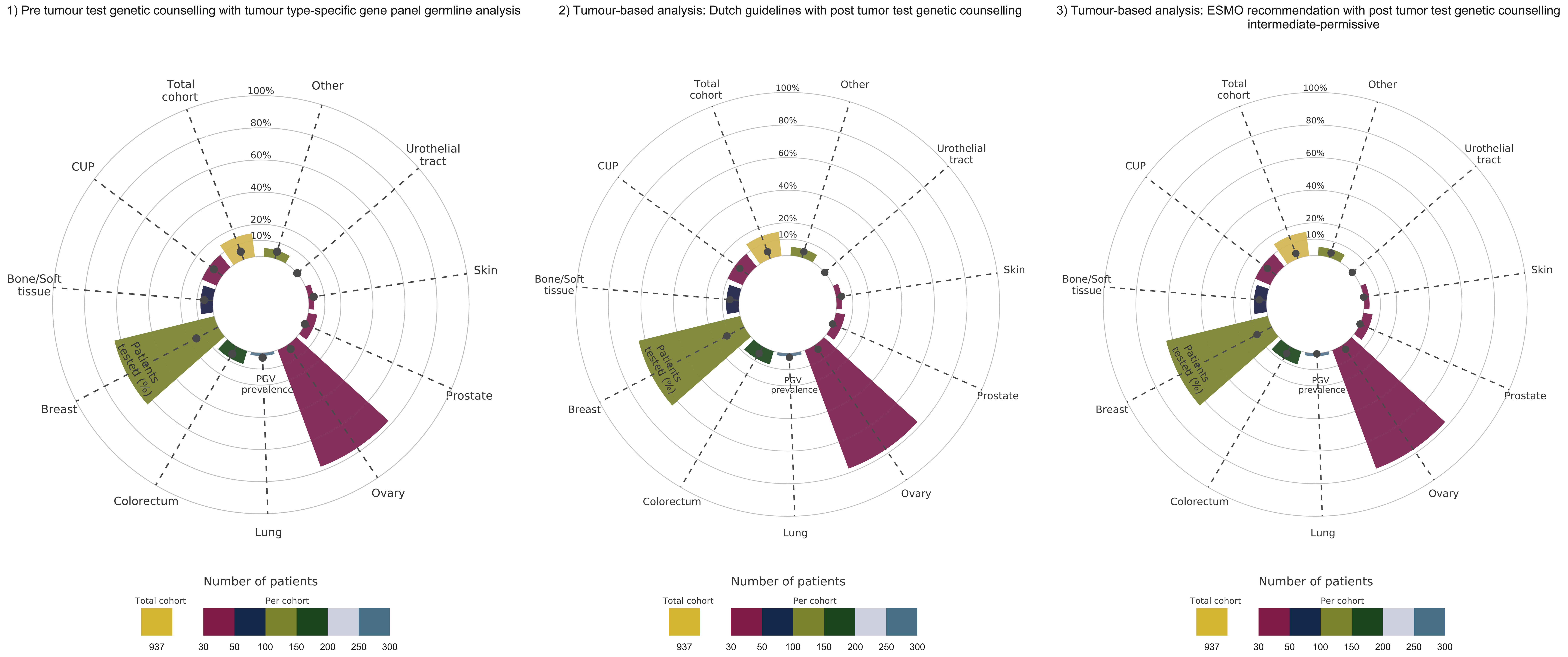

**Supplemental Figure 4.**
Prevalence of pathogenic germline and somatic variants per primary tumour location using the strategy as indicated.
Prevalence of pathogenic germline and somatic variants detected with the GS per primary tumour location/type using the strategy as indicated. Locations with fewer than 30 patients are grouped in “other”. Data based on reference genome version GRCh37/hg19 and gene reference transcripts as indicated in Supplemental Table 1.

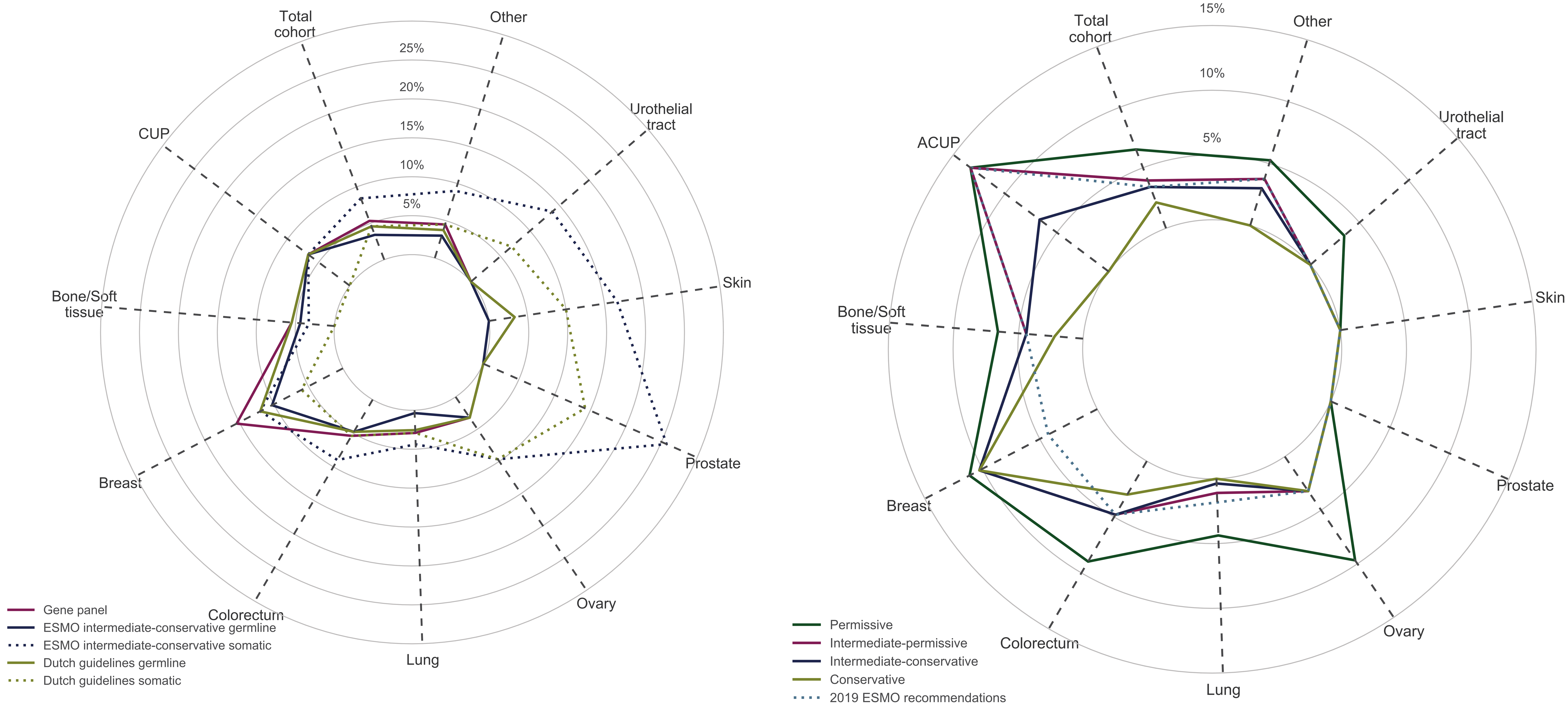
